# The landscape of reported VUS in multi-gene panel and genomic testing: Time for a change

**DOI:** 10.1101/2022.09.21.22279949

**Authors:** Heidi L Rehm, Joseph T Alaimo, Swaroop Aradhya, Pinar Bayrak-Toydemir, Hunter Best, Rhonda Brandon, Jillian G Buchan, Elizabeth C. Chao, Elaine Chen, Jacob Clifford, Ana S A Cohen, Laura K Conlin, Soma Das, Kyle W Davis, Daniela del Gaudio, Florencia Del Viso, Christina DiVincenzo, Marcia Eisenberg, Lucia Guidugli, Monia B Hammer, Steven M Harrison, Kathryn E Hatchell, Lindsay Havens Dyer, Lily U Hoang, James M. Holt, Vaidehi Jobanputra, Izabela D Karbassi, Hutton M Kearney, Melissa A. Kelly, Jacob M. Kelly, Michelle L Kluge, Timothy Komala, Paul Kruszka, Lynette Lau, Matthew S. Lebo, Christian R Marshall, Dianalee McKnight, Kirsty McWalter, Yan Meng, Narasimhan Nagan, Christian S Neckelmann, Nir Neerman, Zhiyv Niu, Vitoria K Paolillo, Sarah A Paolucci, Denise Perry, Tina Pesaran, Kelly Radtke, Kristen J Rasmussen, Kyle Retterer, Carol J Saunders, Elizabeth Spiteri, Christine Stanley, Anna Szuto, Ryan J Taft, Isabelle Thiffault, Brittany C Thomas, Amanda Thomas-Wilson, Erin Thorpe, Timothy J Tidwell, Meghan C Towne, Hana Zouk, the Medical Genome Initiative

## Abstract

**PURPOSE:** Variants of uncertain significance (VUS) are a common result of diagnostic genetic testing and can be difficult to manage with potential misinterpretation and downstream costs, including time investment by clinicians. We investigated the rate of VUS reported on diagnostic testing via multi-gene panels (MGPs) and exome and genome sequencing (ES/GS) to measure the magnitude of uncertain results and explore ways to reduce their potentially detrimental impact.

**METHODS:** Rates of inconclusive results due to VUS were collected from over 1.5 million sequencing test results from 19 clinical laboratories in North America from 2020 - 2021.

**RESULTS:** We found a lower rate of inconclusive test results due to VUSs from ES/GS (22.5%) compared to MGPs (32.6%; p<0.0001). For MGPs, the rate of inconclusive results correlated with panel size. The use of trios reduced inconclusive rates (18.9% vs 27.6%; p<0.001) whereas the use of GS compared to ES had no impact (22.2% vs 22.6%; p=ns).

**CONCLUSION:** The high rate of VUS observed in diagnostic MGP testing warrants examining current variant reporting practices. We propose several approaches to reduce reported VUS rates, while directing clinician resources towards important VUS follow-up.

## Introduction

A growing percentage of the population receives diagnostic genetic testing, yet test reports are often inconclusive due to variants of uncertain significance (VUS). VUS results can pose challenges for patients as well as providers who may lack the technical background and time to investigate and manage these findings (*Gould et al. 2022*). VUS are generally not reported for tests performed for screening purposes (often performed on ostensibly healthy individuals), whereas VUS are typically reported if identified in tests performed on symptomatic individuals for diagnostic purposes. The historical reason for this was to allow ordering providers to conduct appropriate follow-up on VUS with a high likelihood of being causal for a patient. However, the field of genetic testing has evolved over time, with broader indications for usage as well as more genes being interrogated per test, many with low likelihood of identifying causal variants, yet our reporting practices have not similarly evolved to address this change in practice.

The chance of receiving a VUS on a diagnostic genetic test report is influenced by a variety of factors: 1) Clinical presentation and test content - nonspecific clinical features that lead to testing many genes will yield more VUS compared to a highly specific clinical presentation that is correlated with one or small number of genes (e.g., a large MGP test in a patient with muscle weakness vs. testing the *CFTR* gene in a patient with cystic fibrosis).; 2) Ancestry - individuals from underrepresented backgrounds (e.g., African descent) and therefore with less DNA similarity to the “reference genome” and from populations with fewer prior reported pathogenic variants will have more VUS (Gudmundsson et al., 2022); 3) Laboratory reporting practices - laboratory policies typically call for reporting VUS if the test is performed on a symptomatic individual and not reporting VUS if it is a screening test on a healthy individual; and 4) Family sample inclusion - the use of additional family members (e.g., unaffected parents or affected family members) to contextualize the patient’s analysis and either rule out particular variants or provide support for others. It should be noted that exome and genome sequencing (ES/GS) tests interrogate thousands of genes and therefore have the potential to generate the highest rate of VUS. However, ES/GS also utilize different reporting practices and analysis techniques, along with clinical correlation, to decide which variants to report. We wondered whether these differences yield a lower rate of reported VUS for ES/GS, despite the substantially higher number of genes examined.

We sought to explore this question by analyzing aggregate genetic testing results, with the primary goal to understand which test types–MGPs versus ES/GS –lead to more VUS. Given that understanding, we aimed to identify future practices to reduce the burden of VUS on providers and patients. We were able to analyze the effects of test type, clinical indication, and inclusion of parental samples on the rates of inconclusive genetic test results due to VUS.

## Materials and Methods

### LABORATORIES AND TIME PERIOD

Data from diagnostic (symptomatic) testing was collected from 19 clinical laboratories in North America including Ambry Genetics (Aliso Viejo, CA), ARUP Laboratories (Salt Lake City, UT), Children’s Hospital of Philadelphia (Philadelphia, PA), Children’s Mercy Hospital (Kansas City, MO), Fulgent Genetics (Temple City, CA), GeneDx (Gaithersburg, MD), HudsonAlpha Clinical Services Lab (Huntsville, AL), Illumina Clinical Services Laboratory (San Diego, CA), Invitae (San Francisco, CA), Mass General Brigham Laboratory for Molecular Medicine (Cambridge, MA), Mayo Clinic (Rochester, MN), New York Genome Center (New York, NY), Quest Diagnostics (Secaucus, NJ), Rady Children’s Institute for Genomic Medicine (San Diego, CA), Stanford Medicine (Palo Alto, CA), The Hospital for Sick Children (SickKids) Genome Diagnostics Laboratory (Toronto, ON, Canada), The University of Chicago Genetic Services Laboratory (Chicago, IL), University of Washington (Seattle, WA), and Variantyx (Framingham, MA). Results were limited to a two-year period spanning from January 1^st^, 2020 through December 31^st^, 2021. Three laboratories reported all (panel, exome, and genome) test results, nine reported panel and exome, two reported panel and genome, four reported genome only, and one reported panel only, as their exome platform was strictly interpreted based on predefined panels of genes selected at the ordering step (Supplemental Table 1).

### DATA COLLECTION

Deidentified summary data was collected and aggregate statistics were calculated for inconclusive test results with at least one VUS. Inconclusive cases without a VUS (e.g., cases with a single heterozygous variant pathogenic for a recessive disease) were not included in the inconclusive rates. To control for the impact of differences in positive yield (e.g., reducing inconclusive results due to a positive yield increase), the VUS rates were also compared among the combined positive and inconclusive test results from the 17/19 laboratories (97.9% of MGPs and 95.3% of ES/GS) where the rate of positive results with VUS were also provided.

MGP results were grouped by the total number of genes analyzed (2-10, 11-25, 26-50, 51-100, 101-200, >200 genes). ES/GS tests were categorized by exome versus genome and by inclusion of family samples: both parents and the patient (trio test) versus less-than-trio (e.g., patient only, single parent with patient, multiple siblings and patient, or other combinations). For some laboratories, test results were further categorized by disease area across twelve broad indications: cardiovascular, hereditary cancer/cancer syndrome, neurodevelopmental/intellectual disability/autism, neurological/muscular/neuromuscular, retinal disease, hearing loss, renal/gastrointestinal, endocrine, metabolic, dysmorphic/skeletal, hematologic/rheumatologic/immunologic, dermatologic or “other”. The average number of genes for each disease testing area was computed by using the midpoint in the panel range (e.g., 6 for 2-10 genes tested) or 201 for panels >200 genes as a single gene number for each test.

As some panels are derived from exome or genome sequencing as a backbone and then restricted to groups of genes for interpretation, these tests were categorized as “panel tests” as long as the test was limited to interpretation of the panel-defined gene content. Laboratories had the option to further describe VUS results by sub-tiers related to the level of evidence for pathogenicity: VUS-High, VUS-Mid, VUS-Low. However, only two laboratories used sub-tiers. Laboratories were asked to provide race/ethnicity/ancestry information for their aggregate testing population if available (Supplemental Table 2). And finally, laboratories provided diagnostic yield (rate of positive results), though these results were not compared between MGPs and ES/GS given the distinct populations undergoing testing (e.g., MGPs had a high proportion of low yield cancer testing which was not observed in ES/GS).

### DATA QUALITY CONTROL AND EXCLUSIONS

Single-gene test data were collected but excluded from the analysis given that these tests are often performed as follow-up to carrier screening and not offered as diagnostic tests, and this distinction was not tracked in laboratory systems. One laboratory offered custom add-on options for their tests (i.e., a provider could select one or more genes to add to a panel) but these results were not aggregated with the primary panel result; therefore, this data was excluded from analysis. One laboratory offered customizable panel orders but did not track exact panel size and these data were also excluded. Lastly, one laboratory allowed providers to opt-out of VUS reporting for large neurodevelopmental/neurological panels and, as such, cases in which the provider opted out of VUS reporting were excluded. In some cases, if a patient had multiple tests ordered, only the largest panel was counted to avoid double-counting overlapping genes.

### STATISTICAL ANALYSIS

All statistical analyses were performed using Chi-square analysis with Yates correction. A p-value below 0.001 was considered statistically significant and p values down to 0.0001 are displayed.

## Results

Data from 1,512,306 diagnostic MGPs and ES/GS tests were collected from 19 clinical laboratories in North America. The data spanned 1,463,812 MGPs (96.8%), 42,165 ES tests (2.8%) and 6,329 GS tests (0.4%). Race/ancestry/ethnicity data was provided for approximately half of the tests (770,403 individuals) with 59% White, 10% Hispanic, 8% Black, 4% Asian, 12% Mixed/Other and 8% Not Specified (see Supplemental Table 2).

The rate of test results with at least one VUS in the absence of a causal etiology was significantly lower for ES/GS tests (22.5%; 10,933/48,494) than the rate from MGPs (32.6%; 477,617/1,463,812; p<0.0001) (Fig. 1A). To control for differing positive yield, the VUS rates were also compared among the combined positive and inconclusive test results and statistical significance remained. For MGPs, the rate of VUS results correlated with the number of genes, ranging from 6.0% for 287,811 panel tests of 2-10 genes to 76.2% for 84,316 panel tests >200 genes (Fig. 1B,C). Diagnostic yield from ES/GS and MGPs was 17.5% and 10.3% respectively, though the tested populations had noticeably different indications, precluding meaningful comparisons.

**Figure 1.**
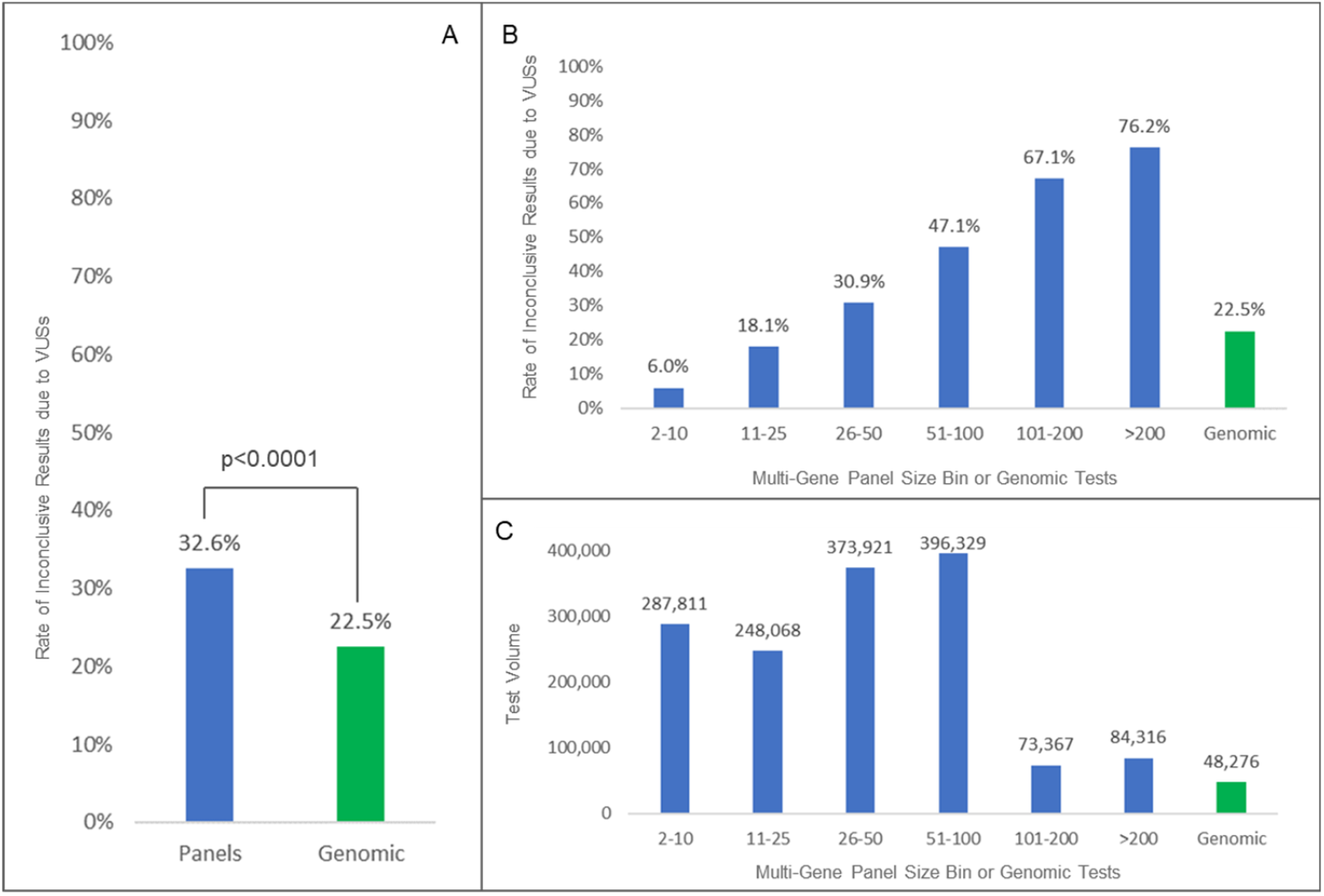
Comparison of Rates of Inconclusive Results due to VUS by Multi-Gene Panel versus ES/GS Testing. Panel A shows a statistically significant reduction in inconclusive rates due to VUS in ES/GS sequencing compared to panels. Panel B shows a breakdown in rates by panel size. Panel C shows test volume for each bin.

All laboratories were able to differentiate ES/GS results by inclusion of family member samples, which we categorized as either a trio (both parents and the patient) versus less-than-trio (only one parent or other combinations of family members without both parents). When examining GS versus ES and trio versus less-than-trio, the use of trios led to significantly lower VUS rates (18.9% vs 27.6%; p<0.0001) (Table 1). There was no difference in the VUS rate when comparing GS vs ES (22.2% vs 22.6%, ns) (Table 1).

**Table 1.**
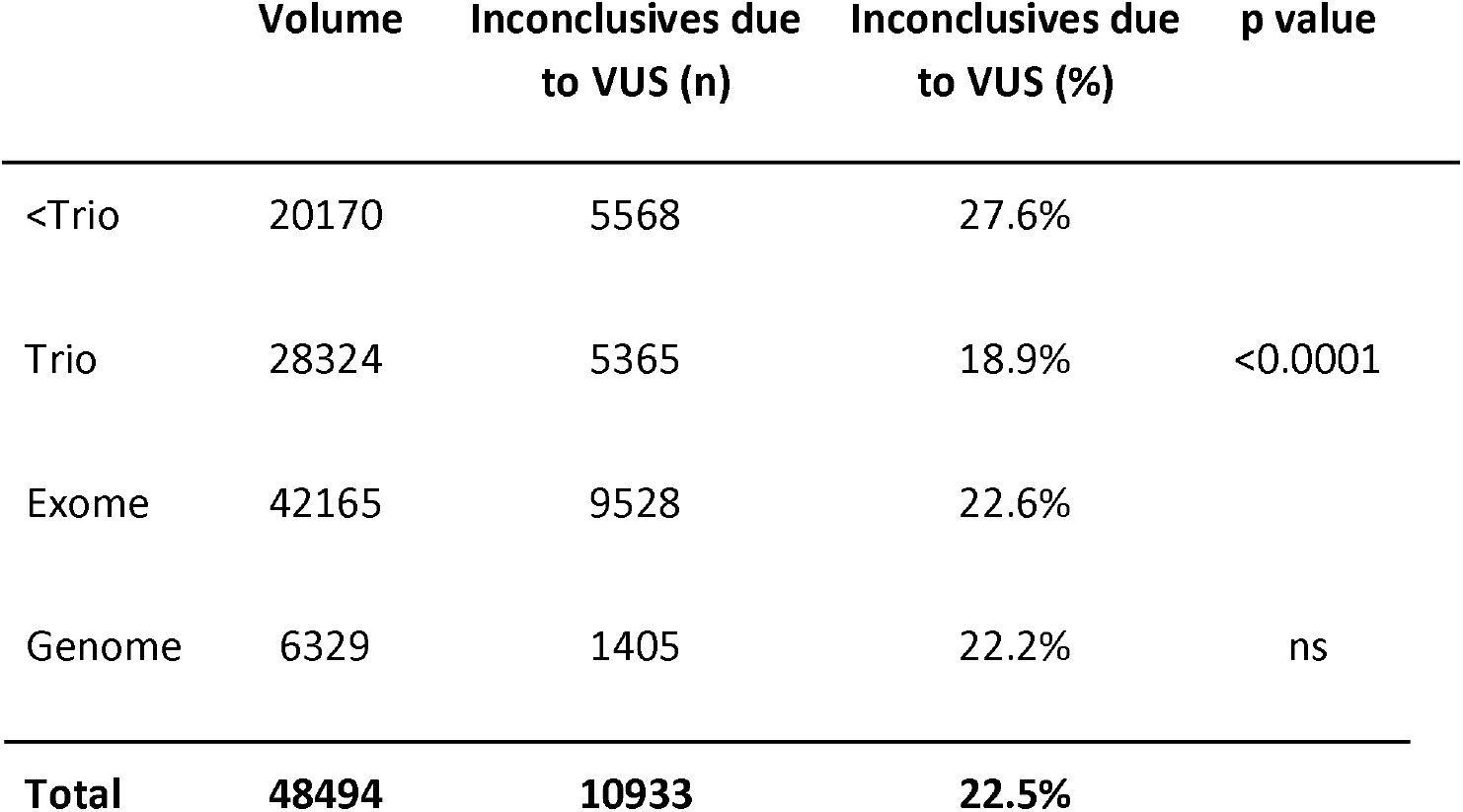
Comparison of ES/GS Inconclusive Rates by Method.

Clinical indications for testing, in the form of disease areas, were specified for 50.2% of MGPs (734,867) and 13.4% of ES/GS tests (6,483). Only six clinical areas had >25 cases and were used to compare differences in VUS rates between MGP and ES/GS tests (Fig. 2; Supp. Table 3). This analysis showed statistically significant lower VUS rates for ES/GS tests compared to MGP tests in cardiovascular, neurologic/muscular, metabolic and neurodevelopmental/intellectual disability/autism. Two disease areas with fewer average numbers of genes tested, hematologic/rheumatologic/immunologic and dysmorphic/skeletal, showed no statistically significant difference in VUS inconclusive rate with ES/GS testing.

**Figure 2.**
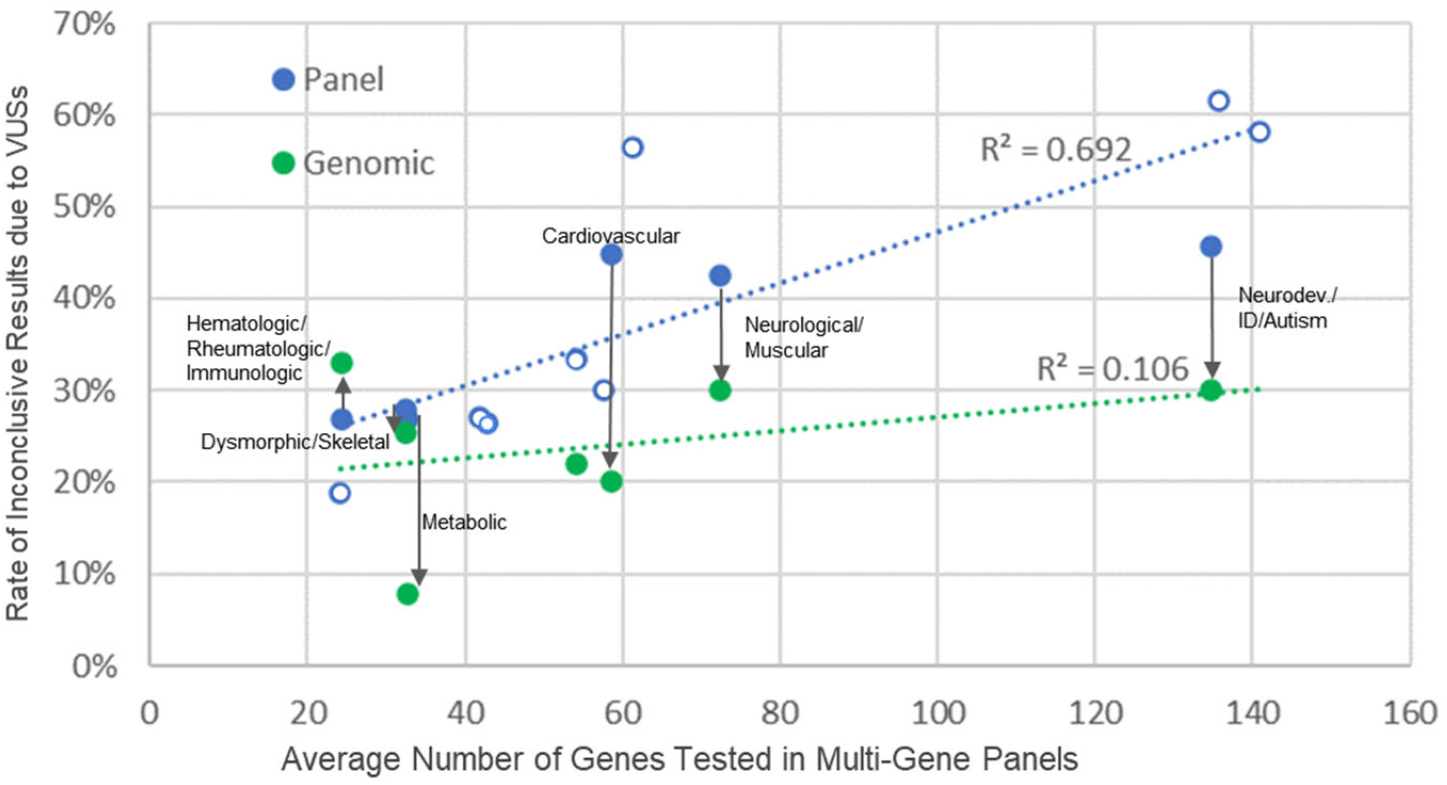
Rate of Inconclusive Results due to VUS by Test Type and Disease Area. Rates generally correlate with multi-gene panel size. Use of genomic testing with trio analysis when available, reduces VUS rate escalation seen with panel size increases. Note: ES/GS results are plotted according to corresponding panel size for the same disease area but actual number of genes analyzed are not captured per ES/GS test. Disease areas (except “not specified”) are labeled when both panel and ES/GS data were available. Open circles represent disease areas with no corresponding ES/GS data. For detailed data for all disease areas, see supplemental Table 3. Supplemental Table 4 includes trio rates for each disease area with comparable data.

Two laboratories (Mass General Brigham LMM and Quest) reported VUS in sub-tiers allowing analysis of the rate of inconclusive results by further subdivisions of VUS evidence. Sub-tier results were mapped to three categories: VUS-High, VUS-Mid, VUS-Low. In this dataset, the sub-tier rate of reported VUS was 22% VUS-High, 56% VUS-Mid, 22% VUS-Low for panels and 40% VUS-High, 60% VUS-Mid, 0% VUS-Low for ES/GS tests (Fig. 3).

**Figure 3.**
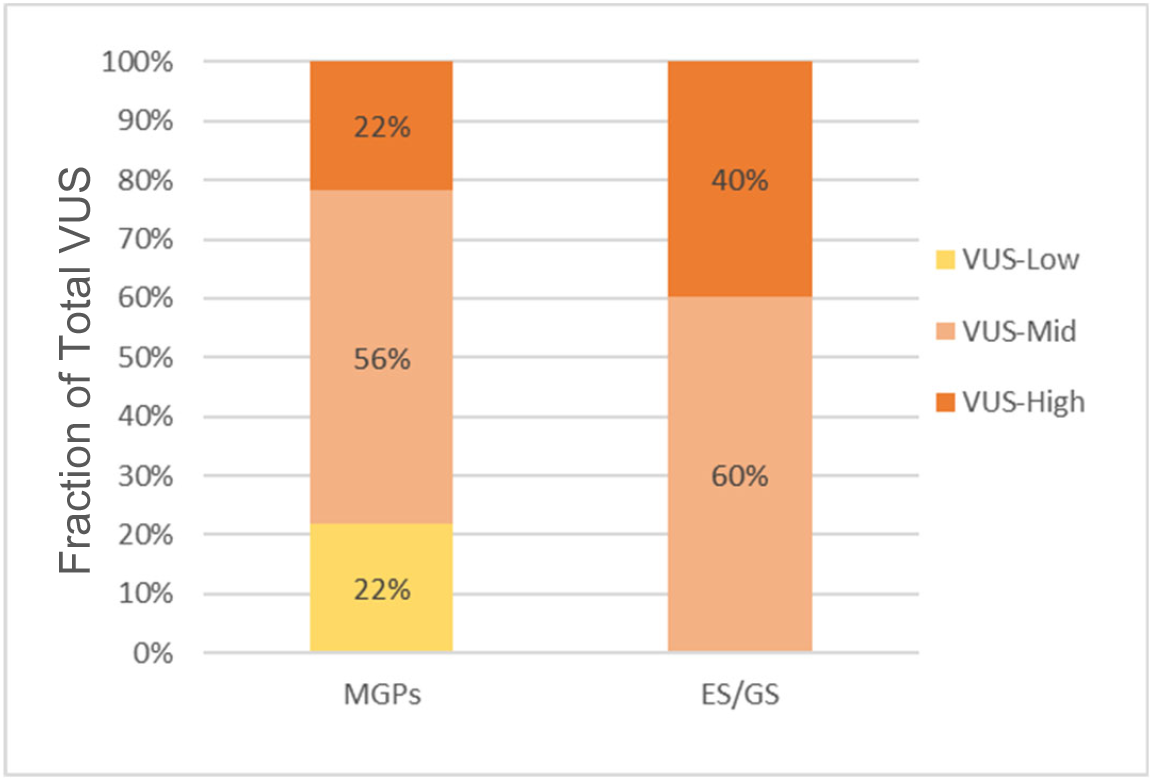
Testing Results Broken down by VUS Sub-tier and Test Type. Two laboratories (Quest and Mass General Brigham) use VUS sub-tiers and these data show the breakdown of VUS reported in MGP and ES/GS testing.

## Discussion

In this study, we observed that exome and genome sequencing (ES/GS) tests demonstrated lower rates of reported VUS compared to multi-gene panel tests (MGPs). This may seem counterintuitive, as ES/GS testing analyzes thousands of genes and millions of variants while MGPs typically analyze a much smaller fraction. However, these results are less surprising when considering the current typical reporting practices of genetic testing laboratories (confirmed to be practiced by all laboratories in this study), which is to report all VUS when performing diagnostic MGP testing. The long-standing rationale in genetic diagnostics has been that reporting VUS in individuals with manifesting disease allows providers the chance to follow up on these results with additional testing that can inform whether or not a variant is pathogenic. For example, a VUS reported in the *GLA* gene can be assessed with α-galactosidase enzyme activity testing in the individual, and these results can provide evidence to inform pathogenicity. Likewise, follow-up family member testing may identify if a variant is inherited or a *de novo occurrence*, if the location of two variants reported for a recessive condition are either in *cis* (same chromosome) or *trans* (opposite chromosomes), or if the variant segregates in other family members with the disease, all of which inform pathogenicity.

However, MGPs have grown to include more and more genes and the range of clinical indications has broadened for each test (e.g., a “pan-cardiac” panel of 100 genes as opposed to a hypertrophic cardiomyopathy panel of 30 genes). While broader panels may be helpful when a clinical diagnosis is uncertain, this results in an increase in VUS rates which are directly correlated with panel size, and an overall weakening in the correlation between the patients’ symptoms (e.g., arrhythmia) and the phenotype associated with any given gene on the panel (e.g., the *MYH7* gene associated with cardiomyopathy). Furthermore, the current ACMG/AMP framework assumes a variant is a VUS until evidence accrues to classify it either as pathogenic/likely pathogenic or benign/likely benign (Richards et al, 2015); yet when testing large panels there is a much lower prior probability that any identified variant is pathogenic.

In ES/GS testing, which would detect the same variants as MGPs and many more, the large quantity of VUS cannot be reported and clinical correlation, segregation analysis and strength of pathogenicity evidence are used to determine which VUS, if any, will be reported. This leads to an overall reduction in the number of VUS reported. In contrast to the extensive clinical information routinely submitted for ES/GS testing to guide analysis and reporting, such information is often absent or very limited when patients are referred for MGP testing. To further aid in VUS reporting and provider follow-up, particularly on MGPs where all VUS are reported, some laboratories use sub-tiers of VUS and indicate the tier on MGP reports or only report the higher evidence tiers for ES/GS testing (Karbassi et al., 2017; Zouk et al., 2021; Baylor Genetics, personal communication). Providers have relayed that these distinctions are helpful to guide which variants to follow-up on and which to ignore.

Limiting VUS results may be desirable for reducing unnecessary follow-up and avoiding mismanagement when VUS are misinterpreted as causal, particularly by providers with less genetics expertise. Indeed, non-cancer patients with VUS have been observed to receive surgical procedures at a higher rate than those without VUS despite many VUS being subsequently reclassified as benign (Walsh et al. 2017). However, one must balance these challenges with the recognition that a subset of VUS, particularly those found in genes well-correlated with specific clinical features, have a higher chance of being found causal, and reporting these VUS along with collaboration with providers is needed to allow evidence to be gathered and tracked over time. It is also important to note that patients may have preferences about the receipt of VUS, in part based upon the balance of potential distress versus future value, as experienced by patients (Skinner et al., 2018; Culver et al., 2013), and providers should ideally engage patients, where possible, in making these decisions.

In general, the high rate of VUS observed in current MGP testing suggests that an examination of current practices is warranted. We consider several approaches that may reduce the overall rate of VUS reported in genetic tests, while at the same time better directing limited provider resources towards important VUS follow-up.

Laboratories could revisit the current practice of MGP panel reporting and consider not reporting all VUS or move those VUS that are less likely causal to a supplemental section of the report. For example, VUS that have very limited pathogenicity evidence (VUS-Low sub-tier), VUS in genes with low correlation between the patient’s phenotype and known gene-disease relationships, or single heterozygous VUS in recessive genes could all be moved to a supplemental reporting section. However, in order to optimize this strategy, ordering clinicians would need to provide more phenotypic data than is often provided for MGP testing, which could be aided by better EHR integration and phenotyping tools provided during test ordering (Son et al, 2018; Owen et al., 2022). In addition, the sub-tier approach will be aided by specific sub-tier guidance anticipated in the release of the next sequence variant classification standards.

Laboratories could offer, or encourage use of, the most focused tests to be used when a patient’s phenotype points to a specific gene list as opposed to unnecessarily ordering very broad MGPs. Providers and patients could also be allowed, and in some cases encouraged, to opt out of VUS reporting, particularly when testing is being performed in individuals with a lower suspicion of a genetic etiology (e.g., most cancer testing). This decision-making could be supported by genetic counseling services or educational decision aids within laboratories or hospital send-out services as well as general physician education approaches (Miller et al, 2014; Hajek et al., 2022). In addition, laboratories could also offer trio-based MGP testing for more severe, non-specific pediatric onset disorders (e.g. epilepsy MGPs) that are more likely to be caused by de novo variation, and highlight for patients and clinicians the added yield from trio-based testing.

### LIMITATIONS

It is possible that differences in the specific populations sent for genetic testing (e.g., suspicion for a genetic etiology, prior testing, specific clinical indications), as well as more detailed differences in laboratory practices for VUS classification and reporting, could impact VUS rates between test types and across disease areas. However, we think any biases introduced by these potential factors are unlikely to impact the main conclusions of the manuscript.

## Conclusion

In summary, this study identified the largest source of VUS results from MGP tests, whereas the use of ES/GS testing reduced the rate of reported VUS. This is best explained by current laboratory practices of reporting all VUS during diagnostic MGP testing compared to the clinical correlation and parental data applied during ES/GS testing, and suggests the approaches utilized in ES/GS testing could be applied more effectively to constrain VUS reporting in MGP testing, particularly for large panels, and other approaches (VUS sub-tiers and report supplements) may benefit both types of testing. As infrastructure and scalable genomic interpretation approaches improve, reducing the complexity and labor involved with genomic interpretation and correlation of genes to phenotypes, as well as more easily enabling the provision of phenotype and family history by clinicians, we anticipate a shift towards ES/GS approaches that can offer the highest sensitivity in testing without added cost compared to MGPs

In addition, it will be prudent for laboratories to provide more education to providers to allow better understanding of the potential clinical significance of a VUS, its likelihood of being classified towards pathogenic or benign, and what specific steps a provider can take to gather evidence to inform the pathogenicity of a variant included on a report. Most importantly, these findings emphasize the critical partnership between providers and laboratories to support the most informative application of genetic and genomic testing to patient care.

## Data Availability

All aggregate data produced in the present study are available upon reasonable request to the authors

## Data Availability

The participating laboratories agreed to provide data on the condition that individual laboratory volumes were not shared. As such, only summary data across laboratories is available and is included in the manuscript and included tables.

## Funding Statement

HR’s participation in this study was funded in part by the National Human Genome Research Institute under award U24HG006834.

## Author Contributions

Conceptualization: C.R.M., C.S., D.dG., D.M., D.P., E.C.C., E.S., H.L.R., H.M.K., H.Z., I.D.K., J.G.B., J.M.H., J.T.A., K.M., K.R., K.R., M.C.T., N.N., P.B-T., R.J.T., S.A., S.D., V.J., Y.M.; Data curation/collection: All authors except N.N., M.E.; Formal analysis/Writing-original draft: H.R.; Writing-review & editing: All authors

## Ethics Declaration

On September 21st, 2022, the Mass General Brigham Human Research Office determined that this project did not meet the criteria for human subject research and therefore did not require IRB approval.

## Conflict of Interest

All authors are or were employed by clinical laboratories offering genetic testing services, as indicated by their affiliations. Additional existing conflicts or those that were relevant at the time of data collection and publication include: Swaroop Aradhya, Elaine Chen, Kathryn E Hatchell, and Dianalee McKnight - Stockholders of Invitae; Christina DiVincenzo, Izabela D Karbassi - Stockholders of Quest Diagnostics; Kyle Retterer - past Stockholder of Sema4 and Opko Health; Kyle W Davis, Nir Neerman, and Christine Stanley - Stockholders of Variantyx; Denise Perry, Ryan Taft, Erin Thorpe, and Brittany Thomas - Stockholders of Illumina, Inc

## Notes

### Competing Interest Statement

All authors are employed by clinical laboratories offering genetic testing services, as indicated by their affiliations. Additional conflicts include:
Swaroop Aradhya, Elaine Chen, Kathryn E Hatchell, and Dianalee McKnight - Stockholders of Invitae;
Christina DiVincenzo, Izabela D Karbassi - Stockholders of Quest Diagnostics; Kyle Retterer - Stockholder of Sema4 and Opko Health; Kyle W Davis, Nir Neerman, and Christine Stanley - Stockholders of Variantyx

### Author Declarations

NOT HUMAN SUBJECT RESEARCH DETERMINATION Date: September 21, 2022 Title of Project: Comparing the rate of uncertainty generated from panels versus genomic sequencing tests Project Lead Name: Heidi Rehm The above referenced project does not meet the criteria for human subject research as defined by Mass General Brigham Human Research Office policies and Health and Human Services regulations set forth in 45 CFR 46. Based on the information you provided this activity is not human subjects research because does not involve human subjects. The project does not require IRB approval. This NHSR activity is not applicable for Clinicaltrials.gov registration. Please retain a copy of this letter in your project file. xPlease feel free to contact our office directly with any questions related to this determination. Sincerely, Ben McGill, MS, CIP Expedited Specialist II, Human Research Affairs

### Summary of Updates

The manuscript was revised in order to submit to a second journal.

