## Supplemental Material for "The landscape of reported VUS in multi-gene panel and genomic testing: Time for a change"

**Supplemental Table 1**

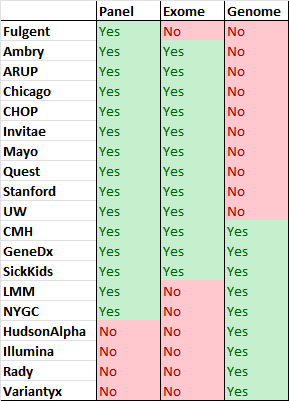

**Supplemental Table 2**

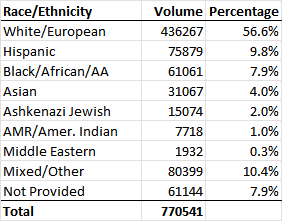

**Supplemental Table 3**

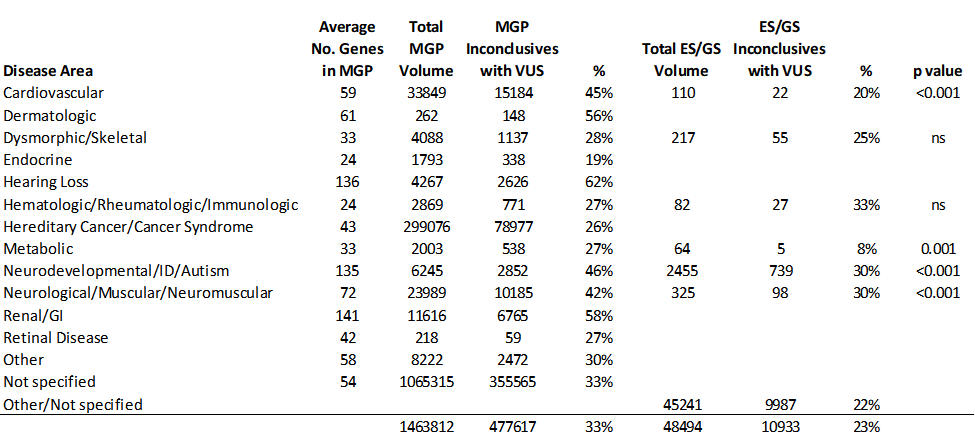

**Supplemental Table 4**

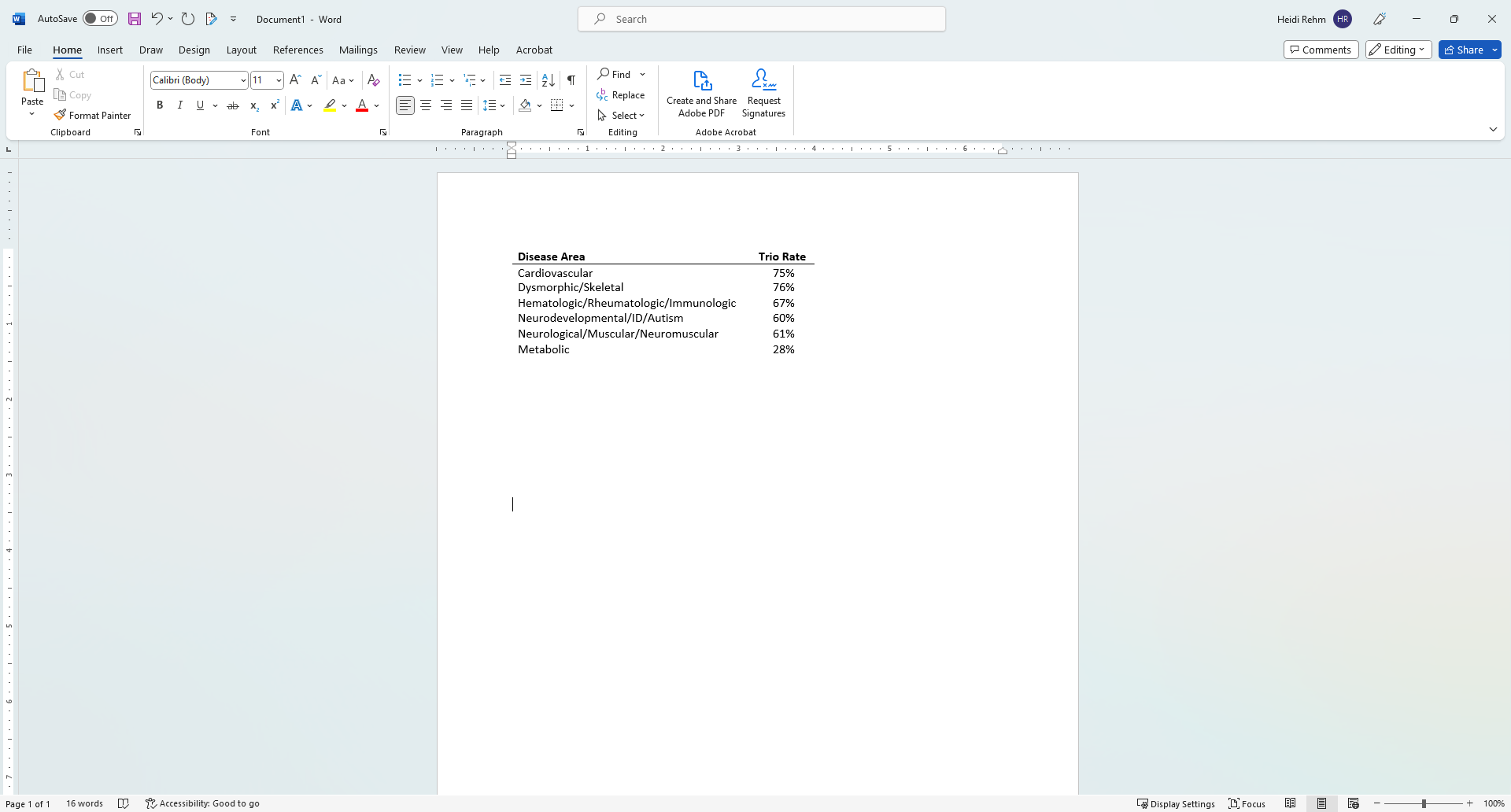

**Data Collection Instructions Provided to Laboratories**

**Duration:** Limit analysis to 2 years: 1/1/2020 - 12/31/21

**Testing context:**

- Diagnostic (symptomatic) germline sequencing testing only (NGS or Sanger)
- Exclude somatic, carrier, population screening, familial variant, genotyping, or any testing that does not report VUS

**Panels:**

- Include panels where analysis is limited to panel genes only
- Exclude “exome slice” type of analyses where analysis may reflex to a wider examination of genes or is customized from entire genome by ordering provider
- Assumption: All variants in a panel are classified and VUS and above reported; if your lab reports fewer variants than this, please include those methods when sharing your data (e.g., do you exclude VUS-Favor Benign?, do you restrict reporting to only variants with a good match to phenotype?, etc)
- Panel size groupings for analysis (you may optionally include the exact panel sizes for each test):
  - 1
  - 2-10
  - 11-25
  - 26-50
  - 51-100
  - 101-200
  - >200
- Disease areas (you may optionally include the exact test name for each test):
  - Cardiovascular
  - Hereditary Cancer/Cancer Syndrome
  - Neurodevelopmental/ID/Autism
  - Neurological/Muscular/Neuromuscular
  - Retinal Disease
  - Hearing Loss
  - Renal/GI
  - Endocrine
  - Metabolic
  - Dysmorphic/Skeletal
  - Hematologic/Rheumatologic/Immunologic
  - Dermatologic
  - Other

**Genome and Exome:**

- Include only genomic testing that makes it clear to the provider that the scope of analysis includes all genes
- Exclude panel testing performed on a exome/genome backbone for lab workflow only if reporting is restricted to the panel
- Separate data by exome vs genome
- Separate cases by trio status:
  - "at least trio” (e.g., proband plus both parents, quads with both parents, etc)
  - “less than trio” (e.g., proband only or proband plus other familial members without both parents)

**Primary data collection variables:**

- Number of Inconclusive cases with at least one VUS
  - Exclude “Positive” cases where a diagnosis was identified but additional VUS were also reported from this category, but we have a separate category (positives with VUS) to report on this
  - Include cases/VUS regardless of whether they are VUS in GUSs vs VUS in known genes
  - Ideally include only inconclusives due to, at least in part, VUS, not those that are solely inconclusive due to other reasons without any VUS (e.g., unclear gene-phenotype match, single het recessive cases). If it is not possible to separate, then note this in your caveats
  - Do not consider secondary findings when assessing the result (e.g., if the diagnostic test result is inconclusive due to a VUS, but you also have a pathogenic secondary finding reported, treat the cases as inconclusive due to the VUS) - there is a separate field where groups can optionally report the number of cases with a secondary/incidental finding (you could add a note on the site-specific details tab to indicate the range of secondary/incidental findings reported by your lab) - you do not need to delineate what the SF/IF cases had in terms of their dx result
- Results should be based on initial reports/classifications (ignore reclassifications); if not possible, explain in caveats
- Optional data collection for non-case centric data:
  - Total VUS reported in panels vs exome and/or genome
    - We prefer that this be total VUS ‘observations’ such that if a given VUS was observed in 10 cases, you would count that 10 times. If you cannot count VUS observations, and can only count unique VUS then please describe that in site-specific details
- Optional data collection for VUS subcategory
  - Written as VUS-FP, VUS-Equivocal, VUS-FB but these can map to whatever terms you use in your lab; you can use only those categories you use (VUS-Suspicious/Favor Path only for example)

**Optional data cleaning:**

- You may exclude cases where a VUS was included in the genome/exome report only because it was previously reported by a panel test

**Race/ethnicity/ancestry:**

- If available, please include the general breakdown of race/ethnicity/ancestry, as you collect it, for your laboratory in the site-specific tab
- You may optionally segregate your data by race/ethnicity/ancestry (simply add more lines in the spreadsheet and separate data by race/ethnicity/ancestry (add column for race/ethnicity/ancestry field)

**Site-specific data to collect:**

- Lab Name
- Primary contact for project
- Lab website URL
- Race/ethnicity/ancestry breakdown of your test population if collected
- Please describe any reporting methods or practices that are relevant to this project and your site:
- Please include any VUS reporting policies/practices for your lab. An example shown below:
  - If a case is positive with a Path variant that supports dx we don’t report VUS
  - We don’t report syn VUS if the variant is not predicted to affect splicing.
  - We only report +/- 10bp into the intron
- Describe any caveats of your data collection:
- Please list individuals who contributed substantially to the oversight, data collection, analysis or manuscript writing for this project
  - Note, this can be updated later if things change
